# Off-season RSV epidemics in Australia after easing of COVID-19 restrictions

**DOI:** 10.1101/2021.07.21.21260810

**Authors:** John-Sebastian Eden, Chisha Sikazwe, Ruopeng Xie, Yi-Mo Deng, Sheena G. Sullivan, Alice Michie, Avram Levy, Elena Cutmore, Christopher C Blyth, Philip N Britton, Nigel Crawford, Xiaomin Dong, Dominic E. Dwyer, Kimberly M. Edwards, Bethany A. Horsburgh, David Foley, Karina Kennedy, Cara Minney-Smith, David Speers, Rachel L. Tulloch, Edward C. Holmes, Vijaykrishna Dhanasekaran, David W. Smith, Jen Kok, Ian G. Barr, Australian RSV study group

## Abstract

Human respiratory syncytial virus (RSV) is an important cause of acute respiratory infection (ARI) with the most severe disease in the young and elderly^1,2^. Non-pharmaceutical interventions (NPIs) and travel restrictions for controlling COVID-19 have impacted the circulation of most respiratory viruses including RSV globally, particularly in Australia, where during 2020 the normal winter epidemics were notably absent^3–6^. However, in late 2020, unprecedented widespread RSV outbreaks occurred, beginning in spring, and extending into summer across two widely separated states of Australia, Western Australia (WA) and New South Wales (NSW) including the Australian Capital Territory (ACT). Genome sequencing revealed a significant reduction in RSV genetic diversity following COVID-19 emergence except for two genetically distinct RSV-A clades. These clades circulated cryptically, likely localized for several months prior to an epidemic surge in cases upon relaxation of COVID-19 control measures. The NSW/ACT clade subsequently spread to the neighbouring state of Victoria (VIC) and caused extensive outbreaks and hospitalisations in early 2021. These findings highlight the need for continued surveillance and sequencing of RSV and other respiratory viruses during and after the COVID-19 pandemic as mitigation measures introduced may result in unusual seasonality, along with larger or more severe outbreaks in the future.

## Main

Each year RSV causes an estimated 3.2 million hospital admissions and 118,200 deaths in children under five years of age, predominantly in low- and middle-income countries^7^. While this burden is greatest in the young, RSV is clinically significant for all age groups, as re-infection can occur throughout life^8^. The elderly and immunocompromised are particularly at risk of severe infection with intensive care admission and mortality rates similar to that of influenza, posing a considerable threat to residents of long-term care facilities^9,10^. RSV causes seasonal epidemics in both tropical and temperate regions of the world^11^. In Australia, most temperate regions experience seasonal RSV outbreaks during the autumn and winter, often peaking in June-July^12^ and usually preceding the influenza season^13^. In the more tropical northern parts of Australia, RSV activity correlates with the rainfall and humidity patterns of the rainy season from December to March^14^.

NPIs to limit the spread of severe acute respiratory syndrome coronavirus 2 (SARS-CoV-2) have disrupted the typical seasonality of other common respiratory pathogens in many countries^15,16^. Australia’s initial SARS-CoV-2 epidemic was effectively controlled by NPIs^17^, and those same restrictions also suppressed seasonal respiratory virus circulation, particularly for influenza virus and RSV, as the usual winter epidemics were notably absent during 2020^3–5^. While the control measures of each Australian state and territory varied in stringency and duration^18^, all occurred throughout the usual peak of RSV seasonal activity. Interestingly, the impact of NPIs was not consistent across all the common respiratory viruses: rhinoviruses and, to a lesser extent, adenoviruses continued to circulate in Australia during this pandemic period after an initial disruption to usual circulation^5^. The suppression of influenza and RSV activity during the COVID-19 pandemic in the southern hemisphere was also seen in South Africa and New Zealand^19^, where similarly following an initial reduction in circulation, RSV activity rebounded in late 2020^20^ and early 2021, respectively. Marked reductions in RSV activity have also been seen in the northern hemisphere since early 2020, although some European countries^21^ and US states^22^ have recently reported out-of-season spikes in RSV activity in early-mid 2021. There has also been a dearth of RSV sequences submitted to public databases since early 2020, presumably a reflection of the lack of RSV circulation and a focus on SARS-CoV-2 sequencing.

In late 2020, severe out-of-season RSV outbreaks occurred in several Australian states and territories beginning in New South Wales and the Australian Capital Territory (NSW/ACT) and Western Australia (WA). This was followed by outbreaks in Victoria (VIC) throughout the summer in early 2021. To understand the change in seasonal prevalence of RSV in Australia we examined RSV testing data from January 2017 to March 2021, comparing the proportion positive and overall testing capacity in NSW, WA, and VIC (Figures 1 & S1). Before 2020, RSV activity consistently began during mid-autumn (April-May) and normally persisted for six months with an epidemic peak in the middle of the Australian winter (middle of July, weeks 27 to 29). In contrast, RSV activity in 2020 occurred between six and nine months later than historically observed, and at the peak of RSV activity across each state, laboratory-confirmed RSV positivity rates were considerably higher than those of the previous three seasons (Figure 1).

**Figure 1.**
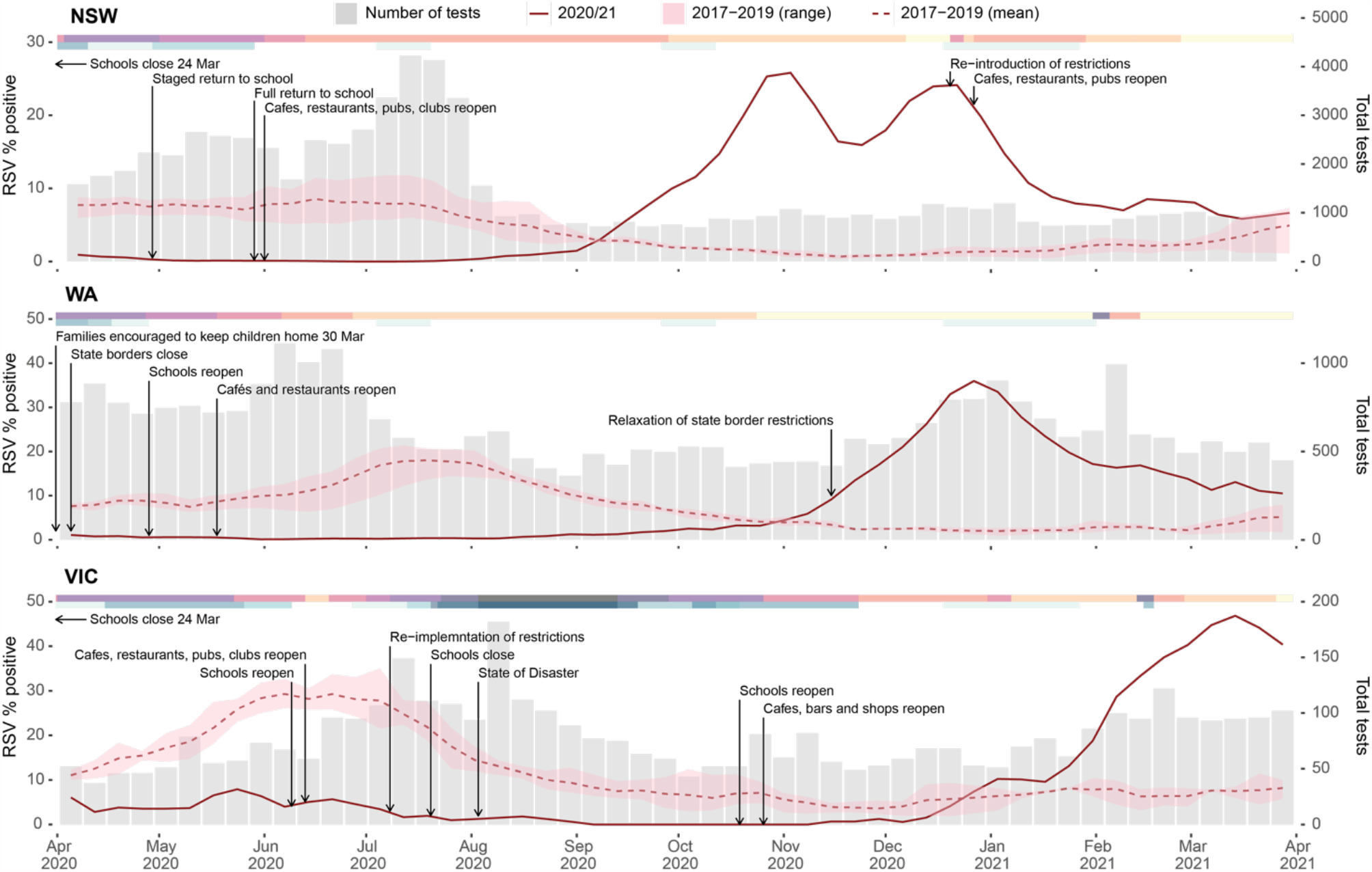
The epidemiology of RSV detections in three Australian states. Laboratory testing for RSV in 2020 as weekly percent positive (red line, left y-axis) and as total number of tests performed (grey bars, right y-axis). In each panel, the dashed red line represents average monthly RSV percent positive over the three previous seasons, and corresponding red shading represents minimum and maximum weekly percent positive. Pink-shaded bars across the top of each plot indicate the severity of pandemic restrictions, with darker colours indicative of greater stringency. Blue bars across the top of each plot indicate the periods during which students did not attend school either due to pandemic restrictions or school holiday periods, with darker colours indicative of more stringent school restrictions.

In Australia, the suppression of RSV activity in early 2020 coincided with restrictions in response to increasing community cases of COVID-19 (Figure 1). During March 2020, individual Australian state and territory governments implemented a range of NPIs, which included limits on international arrivals and strict quarantine requirements (for a minimum of 14 days), internal border closures, social distancing, school closures or encouraging parents to keep their children home, and hygiene protocols to minimise SARS-CoV-2 transmission. Importantly, and most relevant for RSV, childcare centres mostly remained open during these restriction periods. Prior to the implementation of these control measures, a gradual increase in RSV activity was observed in early 2020 across all three states with laboratory test RSV positivity rates being similar to monthly averages over the previous three seasons (Figure S1). The introduction of COVID-19 control measures preceded the rapid decline in RSV incidence in each state. Examination of sentinel hospital records for bronchiolitis by ICD-10 Australian modification (AM) codes (including both RSV-confirmed bronchiolitis and bronchoiolitis of unknown cause) showed a decline that mirrored the decrease in laboratory-confirmed RSV (Figure S1). The subsequent RSV epidemics in late 2020 and early 2021 resulted in test positivity and ICD-10 AM admission levels equivalent to or exceeding the normal winter seasonal RSV activity seen in any of the previous three years. For NSW, the epidemic began in September 2020 with bimodal peaks in activity in mid-November (reaching 26% positivity rate) and early January (reaching 24% positivity rate) (Figure 1). The dual peaks likely reflect inconsistent testing over the Christmas holiday period, as the peak in bronchiolitis hospitalisations in NSW occurred between late December and early January and coincided with this period (Figure 1). Furthermore, a December 2020 peak was also observed in the ACT, where 46% of tests were positive (data not shown). For WA, the RSV epidemic began in late September and peaked in December at 37% positivity, with a matching peak in bronchiolitis hospitalisations (Figure S1).

Over the course of the pandemic, the stringency of COVID-19 restrictions has varied across the different states and territories. In VIC, a second wave of COVID-19 from July to August 2020 necessitated a longer SARS-CoV-2 control period, which likely contributed to a three-month delay in the onset of RSV activity relative to epidemic outbreaks in NSW/ACT and WA. RSV activity in VIC began in early January 2021 and then peaked in early March that year with 48% of tests positive (Figure 1). In each state, the peaks in RSV activity occurred a few months after the relaxation of COVID-19 restrictions (Figure 1).

Whole-genome sequencing was performed on RSV positive specimens collected before (July 2017 to March 2020), during and after (April 2020 to March 2021) the implementation of COVID-19 restrictions in NSW (n=253), ACT (n=47), WA (n=216), and VIC (n=178) (Figure 1C). The samples were mostly collected from young children (median ages between 0.78 -2.34 years for the different states), although all age groups including adults and the elderly were represented. Sampling of gender was even and included geographically diverse locations (Figures S2 & S3). Historically in Australia, both RSV A & B subtypes have co-circulated in shifting but relatively even prevalence^23,24^. This trend continued in the pre-COVID-19 period, where RSV-A comprised between 45% to 79% of cases. However, from late 2020 to early 2021 there was an overwhelming predominance of the RSV-A subtype (>95% for all states). This suggested that RSV-A viruses were responsible for both the NSW/ACT and WA outbreaks in late 2020, as well as, the surge in RSV activity in VIC seen in early 2021.

Phylogenetic analysis of all the available RSV-A genomes revealed that the Australian RSV-A viruses belonged to the ON1-like genotype first reported in Canada in December 2010^25^. These viruses have since become globally predominant and have frequently been re-introduced into Australia^23,26^. Indeed, prior to the emergence of SARS-COV-2 and the related control measures, multiple RSV-A ON-1like sub-lineages co-circulated (Figure 2) with genetic diversity sustained from both endemic and imported sources^23^. While the viruses sampled before March 2020 were well-dispersed amongst those circulating globally, viruses from the post-COVID-19 period formed two geographically distinct monophyletic lineages (Figure 2A). One lineage was associated with cases from NSW and ACT, while the other was associated with cases from WA, and hereafter referred to as the NSW/ACT 2020 and WA 2020 lineages, respectively (Figure 2B). Notably, both genetic lineages were defined by several key non-synonymous changes in the genome. In the WA 2020 lineage, some changes were observed in the glycoprotein (T129I and S174N), nucleocapsid (I104F) and small hydrophobic (H57Q) proteins, while significant non-synonymous variation was observed in the NSW/ACT 2020 virus glycoprotein localised to the C-terminus region (E263Q, L265P, S270P, Y273H, S277P, Y280H, S291P, Y297H, L310P, L314P, and S316P), most of which do not appear to have been previously reported. The RSV-A outbreaks occurred in NSW/ACT and WA during late 2020, and at that time, minimal RSV activity occurred in other Australian states including VIC (Figure 1 & S1). However, genomic analysis showed that the rise in cases in VIC in early 2021 was associated with multiple importations of the NSW/ACT 2020 lineage, with a small number of importations of the WA 2020 lineage, rather than the emergence of another novel lineage (Figure 2A-B).

**Figure 2.**
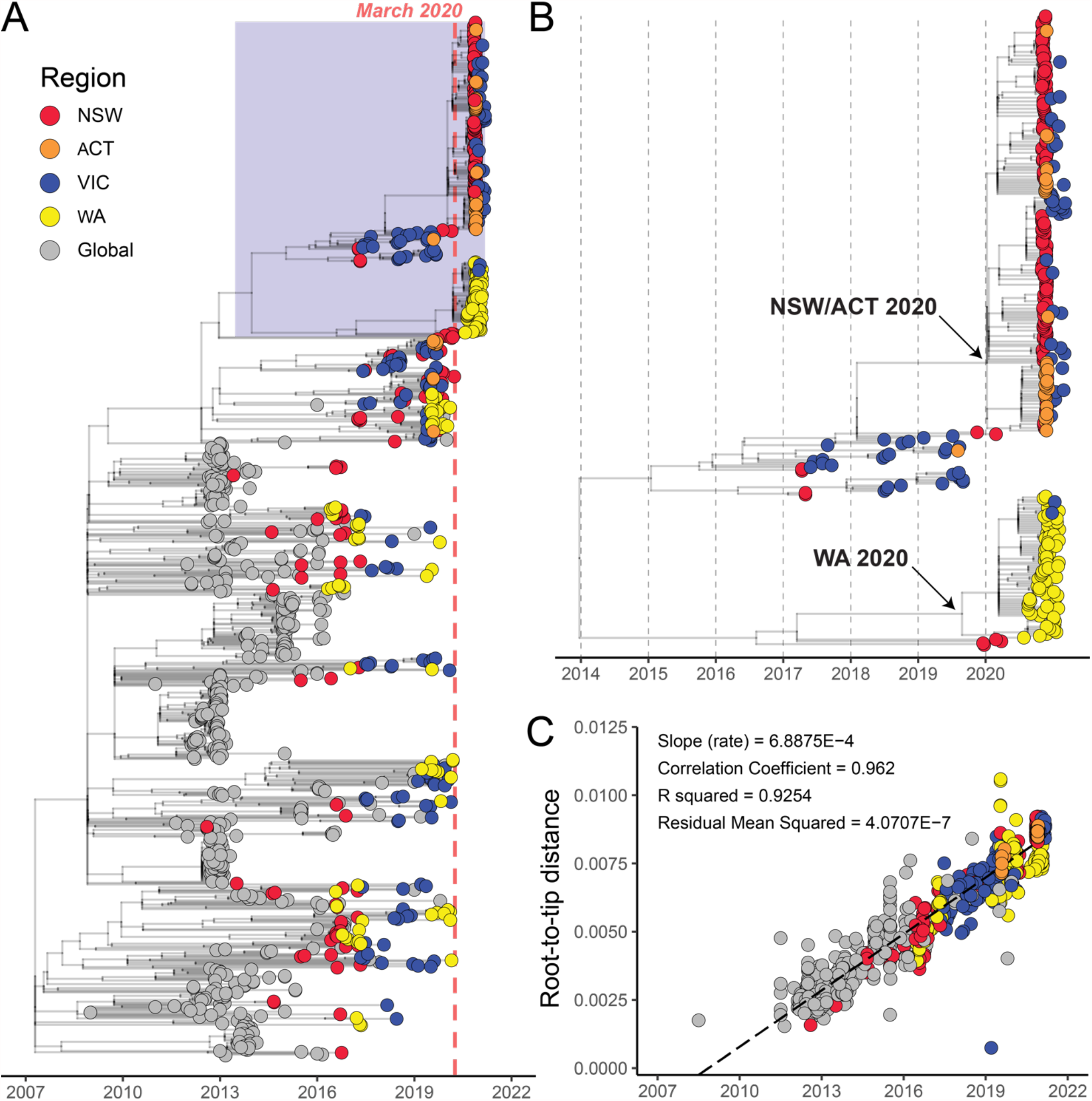
Phylogenetic analysis of global and Australian RSV-A genome sequences. (A) RSV-A genome sequences were aligned with NCBI GenBank reference sequences and analysed using a time-scaled maximum likelihood approach estimated with IQ-TREE focused on recent ON1-like viruses. Australian states and globally-derived sequences are colored according to the key provided. The light red line marks March 2020 and the beginning of extensive COVID-19 related restrictions. The blue shaded box is expanded in panel (B), which is a focused analysis of NSW/ACT and WA 2020 lineages. (C) Temporal signal in RSV-A genomic dataset determined by linear-regression of root-to-tip distance (y-axis) against sample collection date (x-axis).

To maximise spatiotemporal sampling of the RSV-A ON-1like viruses, we expanded our phylogenetic analysis to all globally available RSV glycoprotein (G) sequences (Figure 3A), which are more numerous than published whole RSV genomes. The genome-based phylogeny was comprised of 1130 RSV-A ON1-like genomes (Figure 2A), while a further 1804 sequences were included in the G protein phylogenetic analysis (Figure 3). Despite the additional sequences, the G protein phylogeny found that the viruses from the 2020-2021 epidemics did not cluster with any other viruses sampled nationally or internationally up to date. As such, the initial source of these two novel RSV-A lineages remains undetermined (Figure 3A-B). Similar to the genome-scale analysis, the G protein phylogeny also revealed that the genetic diversity of other ON1-like lineages that co-circulated over the well-sampled period between 2016 and 2020 were absent during these outbreak periods (Figure 3B). We also examined RSV-B diversity, and while the number of detections were low, a similar pattern was observed to RSV-A diversity, whereby previously established lineages were mostly absent, and a single lineage was dominant in the 2020-21 outbreak post-COVID-19 period (Figure S4). Taken together, these results illustrate a remarkable collapse in the genetic diversity of RSV in Australia during the implementation of COVID-19 related restrictions (Figure 3B).

**Figure 3.**
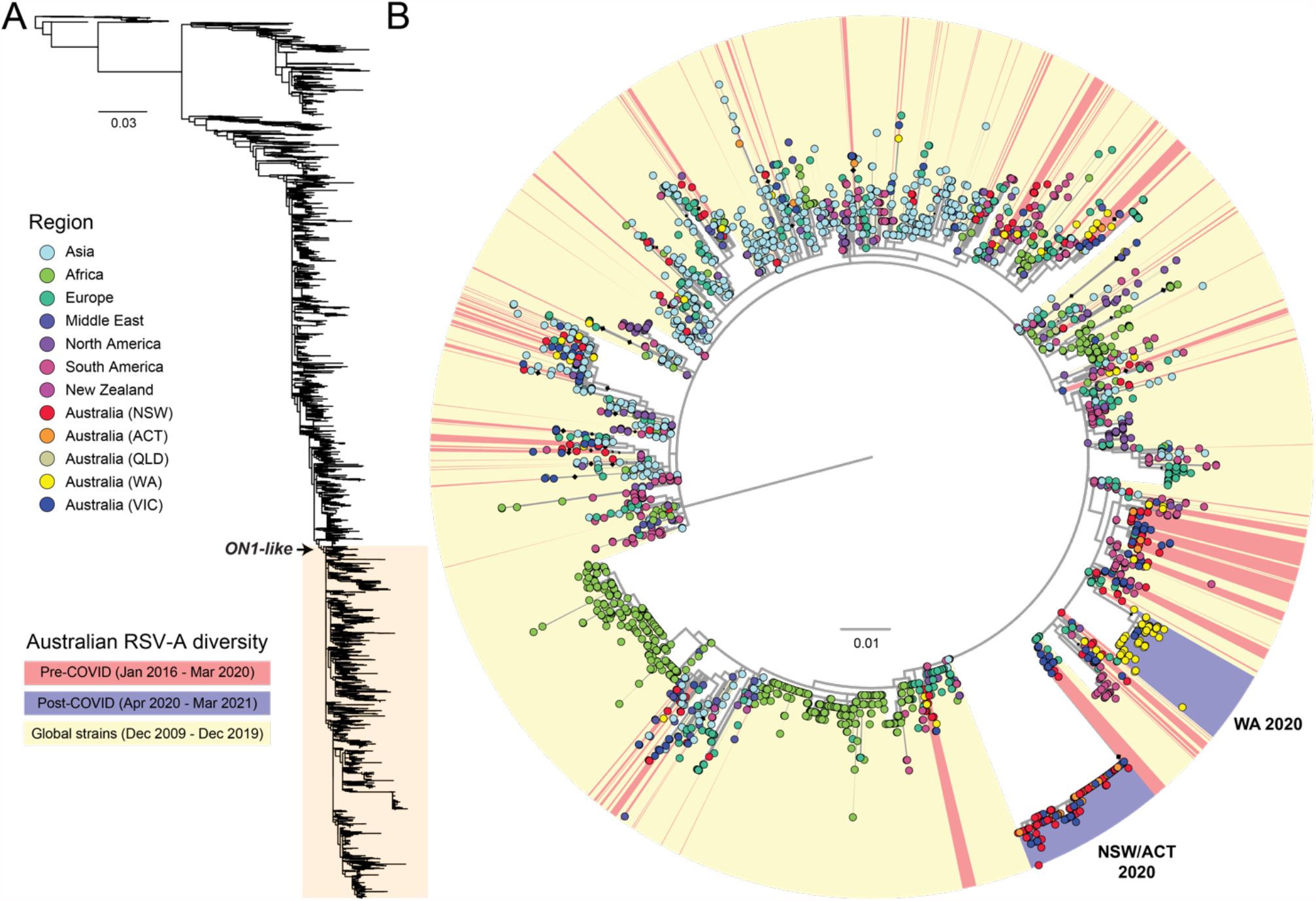
Phylogenetic analysis of global and Australian RSV-A glycoprotein sequences. (A) RSV-A sequences in this study were aligned with all available RSV-A sequences from NCBI GenBank, and the glycoprotein coding region was extracted and sequences less than 300 nt were removed. (B) A detailed examination of recently circulating ON1-like viruses showed pre-COVID-19 lineages (colored red) had collapsed to only two in the post-COVID-19 period (blue) that were associated with outbreaks in NSW/ACT and WA in late 2020. No sequences sourced globally (yellow) were found to be related to the NSW/ACT and WA 2020 lineages suggesting the sources remain unknown. Diamonds at nodes indicate bootstrap support values >70%. Branches are proportional to the number of nucleotide substitutions per site.

The origins of these novel lineages remain unclear. Phylogenetic analysis suggests there was sufficient genetic diversity within the outbreak samples to indicate circulation of these viruses in NSW/ACT and WA prior to the release of COVID-19 restrictions (Figure 2B). Surveillance data support this inference for the WA 2020 lineage; which was first detected in central and southern non-Metropolitan regional WA from late July to September, and then in the Perth Metropolitan area in October (Figure S3), indicating low-level circulation and rapid spread soon after introduction into areas of high population density, in this case Metropolitan Perth. The genomic data also showed two transcontinental transmissions from WA to VIC after November, at a time when interstate travel bans had eased (Figures 2 & 3). In contrast, the origin and early dissemination of the NSW/ACT 2020 lineage was less certain. Despite the widespread occurrence of outbreaks in NSW and ACT, and the relatively high genetic diversity among the sub-lineages in this outbreak (Figure 2 & 3B), no precursor virus(es) of the same lineage were detected prior to the outbreak onset, despite efforts to identify and sequence cases during the low activity period earlier in 2020. In addition to a lack of RSV positive samples throughout the middle of 2020, our analysis was hampered by an inflated evolutionary rate due to recent sampling^27^, multiple novel non-synonymous substitutions observed in the G protein, and a high amount of rate variation that hampered reliable phylogenetic dating estimates. Therefore, we cannot rule out either hypothesis that the viruses were already present in the country before the start of the COVID-19 pandemic or that there were international incursions that occurred during the middle of 2020 when COVID-19 restrictions were in place.

An examination of RSV diversity in Australia before and after the implementation of COVID-related NPIs using whole genome sequencing has shown a major collapse in circulating lineages observed prior to 2020^23,26^. This also coincided with the emergence of two distinct but phylogenetically related RSV-A lineages associated with the summer outbreaks in NSW/ACT and WA, respectively. Both these lineages were found to seed new outbreaks in VIC during early 2021. Our analysis suggests the cryptic circulation of both lineages, while other RSV-A and -B lineages were largely eliminated through COVID-19 related NPIs. While the estimation of the location of origins of the epidemic was hindered by a paucity of sequence data from other jurisdictions and globally, the genetic diversity observed during these outbreaks strongly indicates a localized undetected circulation prior to the widespread outbreak; showing that travel restrictions and other social distancing measures may have significantly reduced RSV transmission and slowed spread, but were unable to eliminate RSV in metropolitan WA and in NSW, and after a considerable delay a strong out-of-season epidemic occurred.

The near absence and subsequent resurgence of RSV-A in Australia has provided a unique opportunity to increase our understanding about how RSV epidemics occur and to identify measures for better control of RSV and other respiratory viruses in the future. Our study highlights how quickly respiratory pathogens can rebound, even leading to unseasonal epidemics. Delayed or forgone RSV seasons may increase the cohort of young children susceptible to RSV infection and increase the age of first infection leading to more frequent and larger outbreaks of RSV when they do finally occur. Indeed, delaying the age at first infection may be expected to coincide with reduced hospitalisations given this burden is most pronounced in infants less than 6 months old^28^; however, this has not been reflected in bronchiolitis admissions (Figure S1), with peak admissions in WA and VIC higher than in prior seasons. By increasing the pool of susceptible children, including those with underlying risk factors such as congenital heart disease and extreme prematurity or chronic lung disease, outbreaks may also be more severe with regards to hospitalisations and intensive care admissions. Indeed, recent modelling studies predicted delayed and severe RSV outbreaks in the US during winter of 2021-2022^15^ but not early out-of-season outbreaks. It remains to be seen whether these large summer epidemics experienced in NSW/ACT, WA and VIC have sufficiently reduced the susceptible population to result in a smaller than usual winter season in 2021, although early winter data appears to support this premise.

It remains unclear how long it will take for normal winter RSV seasonality to resume. The H1N1 2009 influenza pandemic impacted on respiratory virus circulation for a number of years^29^. The findings from this study exemplify the need to be prepared for the occurrence of large outbreaks of RSV outside of normal seasonal periods and for health systems to be prepared to combat future severe RSV outbreaks. It also raises important questions as to how the epidemiological and evolutionary dynamics of RSV outbreaks might inform the re-emergence of influenza virus, which is still expected, and given the smaller role children play in population-scale transmission, may require re-opening of international borders to import the variants required to effectively seed new local outbreaks. Nonetheless, our study highlights the power of COVID-19-related NPIs to cause immense disruption in seasonal patterns of respiratory virus circulation and evolution. Furthermore, this study provides a timely warning to countries emerging from pandemic restrictions that the burden of disease from other respiratory pathogens that may have all but disappeared, will likely rebound in the near future, possibly at unusual times and with higher magnitude of cases.

## Methods

### RSV surveillance and epidemiology

Respiratory specimen testing with quantitative RT-PCR assays was performed at six sites including i) NSW Health Pathology - Institute of Clinical Pathology and Microbiology Research (ICPMR), Westmead, NSW, ii) The Children’s Hospital at Westmead, NSW, iii) PathWest Laboratory Medicine WA, Perth, WA^30^, iv) The Royal Children’s Hospital, Melbourne, VIC, v) Monash Pathology, Monash Medical Centre, Clayton, VIC and vi) ACT Pathology, Canberra, ACT. NSW Health Pathology - ICPMR and PathWest laboratories are both major diagnostic hubs that provide state-wide testing for respiratory viruses in NSW and WA, respectively, Monash Pathology provides services for all ages for a region of Melbourne, while the Children’s Hospital at Westmead and Royal Children’s Hospitals are major metropolitan hospitals in NSW and VIC, respectively. ACT Pathology provides diagnostic services for all adult and paediatric hospital emergency department presentations and a proportion of outpatient community requests for the ACT. Weekly counts for RSV testing were collated for the period January 2017 to March 2021, and derived from three laboratories: PathWest Laboratory in Perth, WA, NSW Health Pathology - ICPMR in Sydney, NSW, ACT Pathology, Canberra, ACT, and the Bio21 Royal Children’s Hospital in Melbourne, VIC. PathWest, ICPMR and ACT Pathology are public health laboratories testing children and adults across their respective states, whereas Bio21 only provided testing data for children receiving care at the Royal Children’s Hospital in Melbourne, and therefore only includes results for children aged under 18 years. The proportion of tests that were RSV positive was calculated and smoothed using a 3-week, centred moving average. Data were plotted in time series to compare observed RSV activity for April 2020 to March 2021, versus the average for April 2017 to March 2020. Monthly bronchiolitis admissions for the 3 children’s hospitals in Perth, Sydney and Melbourne were collated for the period January 2017 to March 2021. Only admissions with a J21 ICD-10 AM code were considered. Prior work has shown that admissions for bronchiolitis are heavily represented by children aged <2 years. School and other restrictions for each state were collated from media releases and official public health directions issued in each of the three states. In addition, school holiday periods for all years January 2017 to March 2021 were collated to visually assess the role of school holidays as a proxy for student mixing in RSV seasonality. The period between relaxation of restrictions and increased RSV activity was visually compared.

### RSV subtyping and whole genome sequencing

Samples were sequenced from cases collected for routine diagnostic purposes as part of public health responses, and from on-going research studies approved by the local Human Research Ethics Committees of the Royal Children’s Hospital and Western Sydney Local Health District with approval numbers 37185 and LNR/17/WMEAD/128, respectively. Total nucleic acid was extracted from RSV positive respiratory specimens archived at -80°C using high-throughput bead-based protocols. RSV whole genome sequencing (WGS) was conducted using established protocols^24^ for a subset of samples selected to provide temporal and geographical representation of i) the pre-COVID-19 period, inclusive of July 2017 to March 2020, and ii) the post-COVID-19 period, inclusive of April 2020 to March 2021. Briefly, viral cDNA was prepared from extracted nucleic acid using SuperScript IV VILO Master Mix or SuperScript IV (Invitrogen, Carlsbad, CA, USA), followed by RT-PCR amplification of four long overlapping fragments spanning the RSV genome using Platinum SuperFi Master Mix (Invitrogen). The four target amplicons were then combined equally before DNA purification with AMPure XP (Beckman Coulter, Indianapolis, IN, USA). The purified and pooled amplicons were diluted to 0.2 ng/µl and prepared for sequencing using the Nextera XT library preparation kit with v2 indexes (Illumina, San Diego, CA, USA). Multiplexed libraries were then sequenced either on an Illumina iSeq 100 or MiSeq producing at least 200,000 paired end reads (2×150nt) per library. For genome assembly, the sequence reads were QC trimmed using BBDuk v37.98^31^ before *de novo* assembly with MEGAHIT v1.1.3^32^ or reference based assembly with IRMA^33^. To confirm assembly, the trimmed sequence reads were re-mapped onto the draft genome with BBMap v37.98 and visually assessed using the Geneious Prime v.2020.0.3 before the final majority consensus genome was extracted. The sequences generated in this study have been deposited in GISAID (Table S1).

### Phylogenetic analysis

RSV sequences generated in this study were analysed along with reference sequences sourced from the NCBI GenBank database or from the NIAID Virus Pathogen Database and Analysis Resource (ViPR)^34^ at http://www.viprbrc.org/. Specifically, all available full-length genomes and partial G gene sequences (greater than 300 nt) with collection dates were downloaded on 22 March 2021. Multiple sequence alignments were performed independently with MAFFT v.7^35^ and examined using TempEst v.1.5^36^ to identify and exclude excessively divergent sequences in a preliminary maximum likelihood (ML) tree generated in FastTree v.2.1^37^. Phylogenetic relationships of the full-length alignments were inferred using the ML method in IQ-TREE v.2.0^38^ using the best-fit nucleotide substitution model and dated using the Least Squares Dating (LSD) method^39^. Ultrafast bootstrap approximation (UFBoot) and SH-like approximate likelihood ratio test (SH-aLRT) was applied to estimate branch support. G gene phylogenies were estimated using RAxML v.8^40^ using the GTR-𝚪 nucleotide substitution model, with branch support estimated by 1000 bootstrap replicates.

## Data Availability

Sequences generated in this study are available from www.gisaid.org (EpiRSV).

## Acknowledgements

Members of the RSV study group include Annette Alafaci, Ian Carter, Andrew Daly, Michelle Francis, Alison Kesson, Hannah Moore, Christine Ngo & Tyna Tran.

