## Supplementary material for "Off-season RSV epidemics in Australia after easing of COVID-19 restrictions": Figures S1-4 & Table S1

### Off-season RSV epidemics in Australia following COVID-19 restriction easing

### Supplementary material

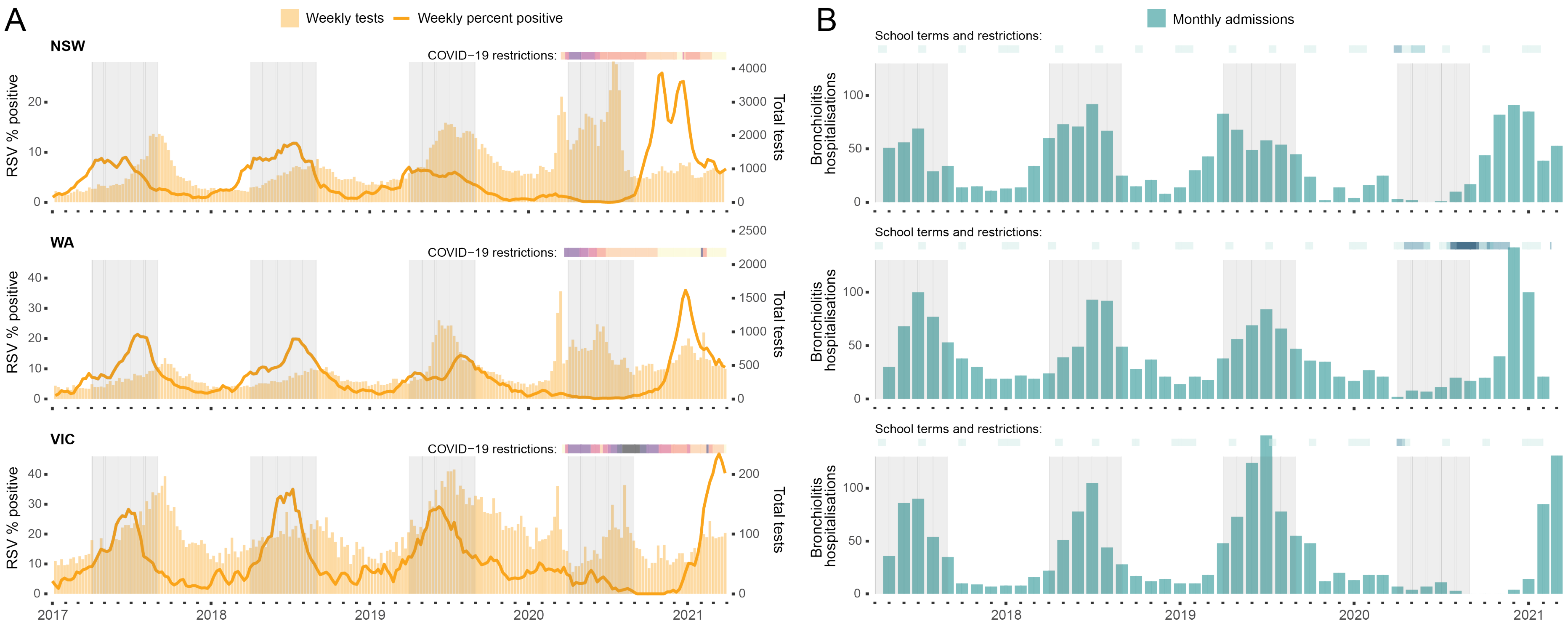


**Figure S1. Time series showing laboratory testing data (A) and RSV bronchiolitis admissions (B) for three Australian states.** Bars at the top of each state indicate COVID-19 restrictions or school holidays/restrictions for each state. For pandemic restrictions, darker colours indicate more severe restrictions. For schools, shading indicates school holiday periods, which darker shading indicates the period during which schools were closed or parents were encouraged to keep their children home. Grey shaded regions indicate the period 1 April to 1 September, which coincides with usual activity of RSV and other respiratory viruses.

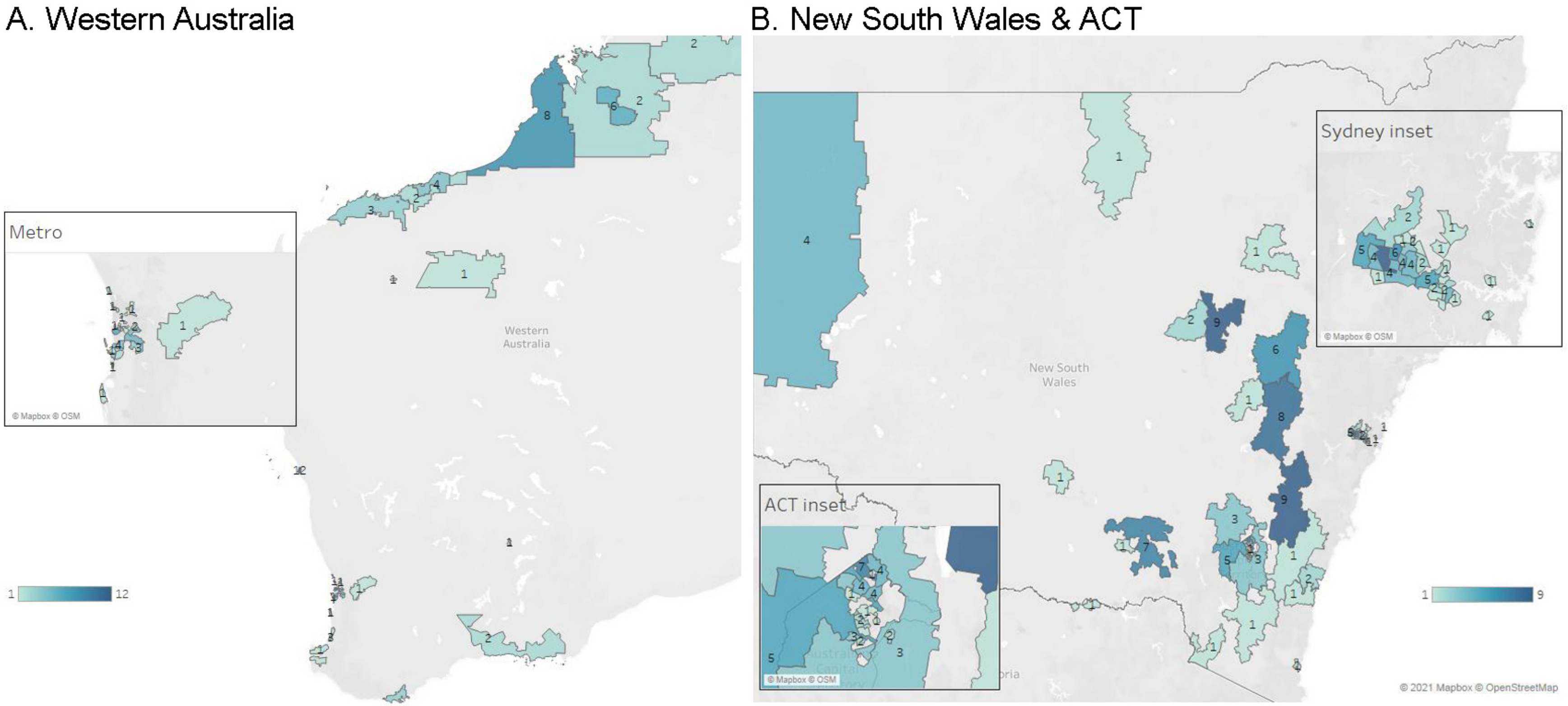


**Figure S2. Geographic distribution of RSV genome sequences in Western Australia (WA) and New South Wales (NSW) and Australian Capital Territory (ACT), Australia**. The number of RSV genomes and their sampling location have been mapped according to postcode and coloured according to the keys provided for each state and territory. Population dense metropolitan areas, including Perth in WA, Sydney in NSW, and Canberra in the ACT are displayed as focused insets.

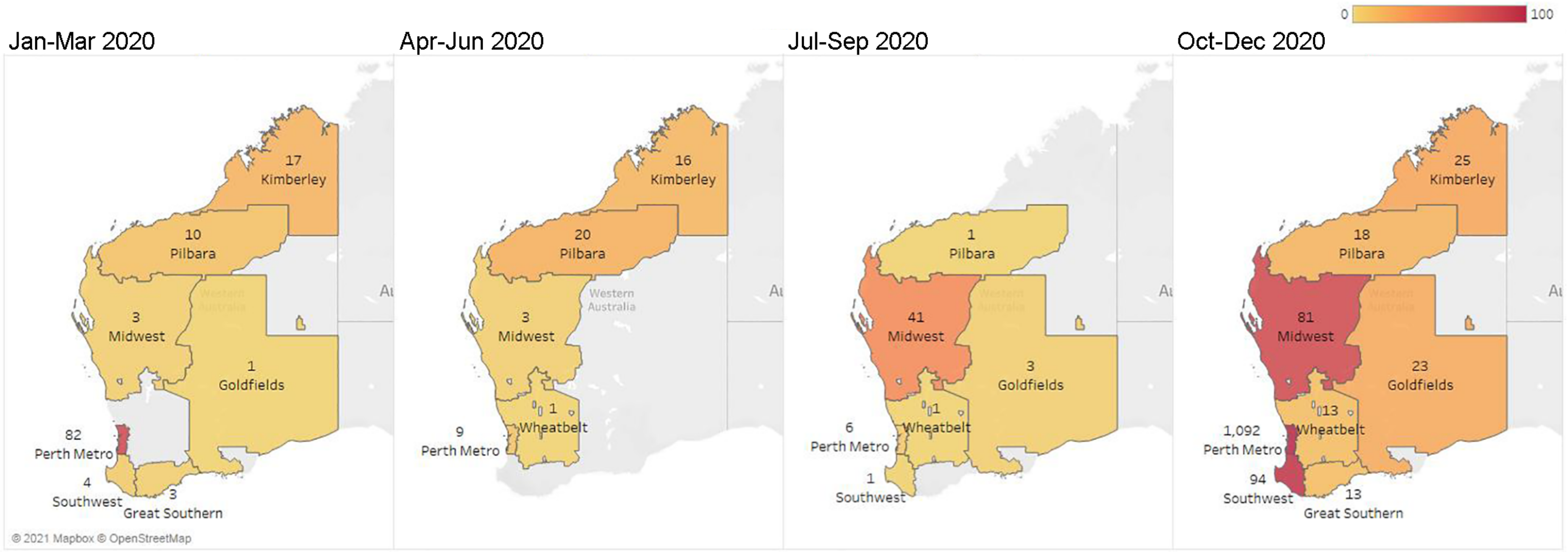


**Figure S3. Quarterly stratified sampling of total number of PCR positive RSV cases that underwent testing at PathWest Laboratory Medicine WA during 2020**. Sampling locations were defined by region and coloured according to the total number of RSV cases within each region per quarter, with the darker colours being representative of increased case numbers.

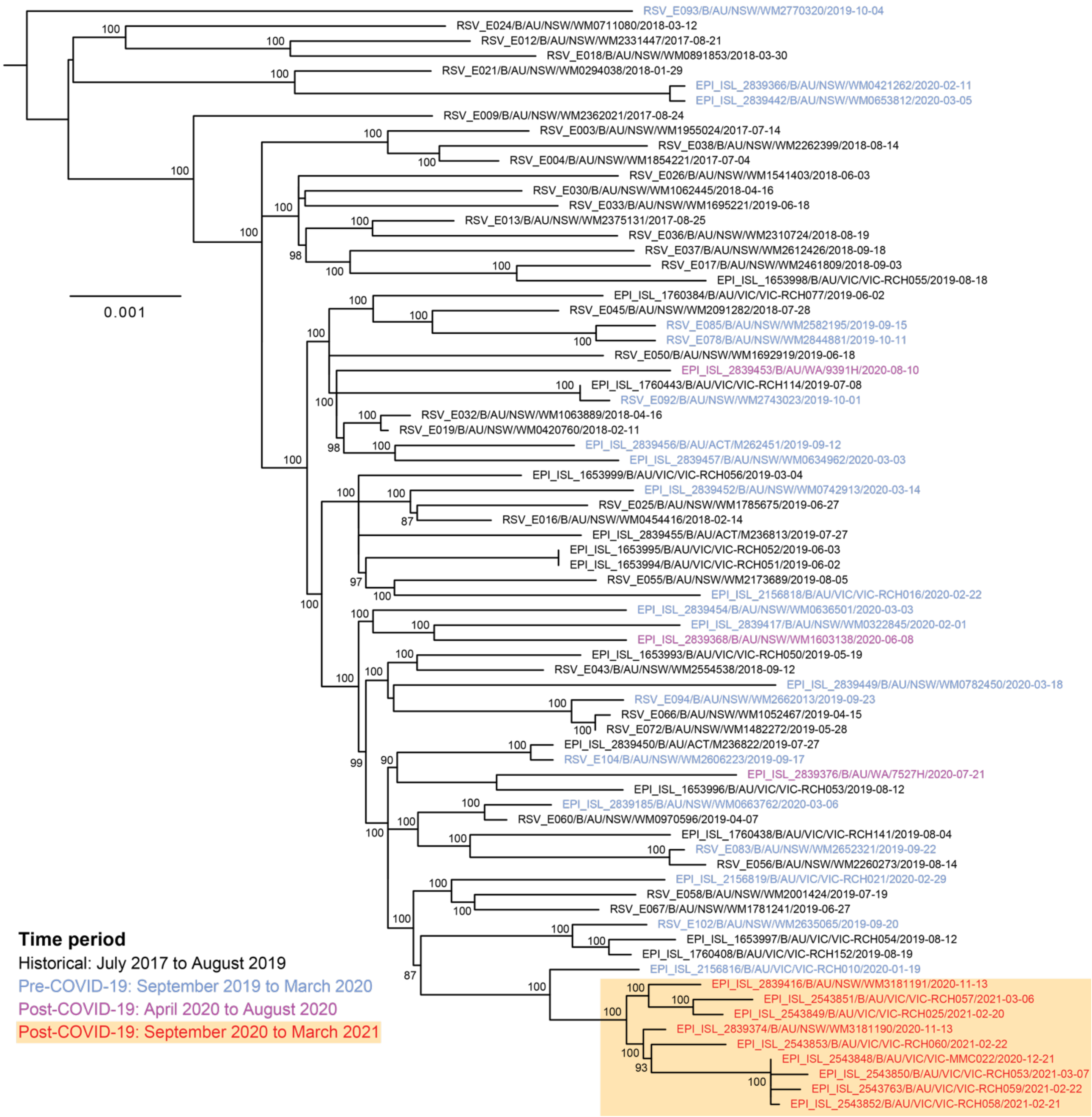


**Figure S4. Phylogenetic analysis of RSV-B diversity in Australia since mid-2017.** RSV-B genome sequences generated in this study (n=74) were aligned and analysed phylogenetically using PhyML. The sequence labels show the accession, subtype, location, strain and collection date, and have been colored according to the period they were collected as per the key provided. Values at nodes indicate bootstrap support values >70%. Branches are proportional to the number of nucleotide substitutions per site. The clade highlighted at the bottom represents a monophyletic lineage that emerged during the outbreak period of late 2020 to early 2021.

**Table S1. Accession numbers for sequences generated in this study (*further accessions pending*)**

| Accession | Virus | Location* | Collection |
|---|---|---|---|
| EPI_ISL_2839213 | hRSV/A/Australia/P511402/2020 | ACT | 2020-11-27 |
| EPI_ISL_2839312 | hRSV/A/Australia/P536277/2020 | ACT | 2020-12-04 |
| EPI_ISL_2839356 | hRSV/A/Australia/P511794/2020 | ACT | 2020-11-27 |
| EPI_ISL_2839310 | hRSV/A/Australia/P520872/2020 | ACT | 2020-11-27 |
| EPI_ISL_2839215 | hRSV/A/Australia/P527755/2020 | ACT | 2020-11-28 |
| EPI_ISL_2839354 | hRSV/A/Australia/P514660/2020 | ACT | 2020-11-20 |
| EPI_ISL_2839351 | hRSV/A/Australia/P510179/2020 | ACT | 2020-11-19 |
| EPI_ISL_2839300 | hRSV/A/Australia/N122610/2020 | ACT | 2020-11-23 |
| EPI_ISL_2839296 | hRSV/A/Australia/P511810/2020 | ACT | 2020-11-27 |
| EPI_ISL_2839227 | hRSV/A/Australia/P527750/2020 | ACT | 2020-11-28 |
| EPI_ISL_2839294 | hRSV/A/Australia/P514373/2020 | ACT | 2020-11-23 |
| EPI_ISL_2839197 | hRSV/A/Australia/P535659/2020 | ACT | 2020-12-04 |
| EPI_ISL_2839451 | hRSV/A/Australia/P522837/2020 | ACT | 2020-11-28 |
| EPI_ISL_2839214 | hRSV/A/Australia/P527905/2020 | ACT | 2020-11-25 |
| EPI_ISL_2839217 | hRSV/A/Australia/P522722/2020 | ACT | 2020-11-29 |
| EPI_ISL_2839231 | hRSV/A/Australia/P508521/2020 | ACT | 2020-11-19 |
| EPI_ISL_2839228 | hRSV/A/Australia/P511846/2020 | ACT | 2020-11-20 |
| EPI_ISL_2839216 | hRSV/A/Australia/P525045/2020 | ACT | 2020-11-30 |
| EPI_ISL_2839218 | hRSV/A/Australia/P533369/2020 | ACT | 2020-12-03 |
| EPI_ISL_2839234 | hRSV/A/Australia/P538685/2020 | ACT | 2020-12-07 |
| EPI_ISL_2839304 | hRSV/A/Australia/P514323/2020 | ACT | 2020-11-23 |
| EPI_ISL_2839302 | hRSV/A/Australia/P507350/2020 | ACT | 2020-11-20 |
| EPI_ISL_2839298 | hRSV/A/Australia/P526371/2020 | ACT | 2020-11-30 |
| EPI_ISL_2839424 | hRSV/A/Australia/P520545/2020 | ACT | 2020-11-29 |
| EPI_ISL_2839308 | hRSV/A/Australia/P507573/2020 | ACT | 2020-11-19 |
| EPI_ISL_2839425 | hRSV/A/Australia/P506175/2020 | ACT | 2020-11-17 |
| EPI_ISL_2839229 | hRSV/A/Australia/P510167/2020 | ACT | 2020-11-23 |
| EPI_ISL_2839230 | hRSV/A/Australia/P520806/2020 | ACT | 2020-11-28 |
| EPI_ISL_2839233 | hRSV/A/Australia/P515852/2020 | ACT | 2020-11-28 |
| EPI_ISL_2839178 | hRSV/A/Australia/P513256/2020 | ACT | 2020-11-22 |
| EPI_ISL_2839232 | hRSV/A/Australia/P525519/2020 | ACT | 2020-11-30 |
| EPI_ISL_2839306 | hRSV/A/Australia/P517150/2020 | ACT | 2020-11-20 |
| EPI_ISL_2839427 | hRSV/A/Australia/P527732/2020 | ACT | 2020-11-28 |
| EPI_ISL_2839190 | hRSV/A/Australia/P520445/2020 | ACT | 2020-11-26 |
| EPI_ISL_2839456 | hRSV/B/Australia/M262451/2019 | ACT | 2019-09-12 |
| EPI_ISL_2839455 | hRSV/B/Australia/M236813/2019 | ACT | 2019-07-27 |
| EPI_ISL_2839450 | hRSV/B/Australia/M236822/2019 | ACT | 2019-07-27 |
| EPI_ISL_2839434 | hRSV/B/Australia/M248046/2019 | ACT | 2019-08-08 |
| EPI_ISL_2839235 | HRSV/A/Australia/WM3181215/2020 | NSW | 2020-11-13 |
| EPI_ISL_2839237 | HRSV/A/Australia/WM3260993/2020 | NSW | 2020-11-21 |

| | | | |
|---|---|---|---|
| EPI_ISL_2839290 | HRSV/A/Australia/WM3200400/2020 | NSW | 2020-11-15 |
| EPI_ISL_2839275 | HRSV/A/Australia/WM3081573/2020 | NSW | 2020-11-03 |
| EPI_ISL_2839256 | HRSV/A/Australia/WM3160671/2020 | NSW | 2020-11-11 |
| EPI_ISL_2839264 | HRSV/A/Australia/WM3090116/2020 | NSW | 2020-11-04 |
| EPI_ISL_2839208 | HRSV/A/Australia/WM3071668/2020 | NSW | 2020-11-02 |
| EPI_ISL_2839279 | HRSV/A/Australia/WM3140467/2020 | NSW | 2020-11-09 |
| EPI_ISL_2839277 | HRSV/A/Australia/WM3120229/2020 | NSW | 2020-11-07 |
| EPI_ISL_2839252 | HRSV/A/Australia/WM3220890/2020 | NSW | 2020-11-17 |
| EPI_ISL_2839212 | HRSV/A/Australia/WM3080869/2020 | NSW | 2020-11-03 |
| EPI_ISL_2839422 | HRSV/A/Australia/WM3110533/2020 | NSW | 2020-11-06 |
| EPI_ISL_2839288 | HRSV/A/Australia/WM3150812/2020 | NSW | 2020-11-10 |
| EPI_ISL_2839266 | HRSV/A/Australia/WM3202125/2020 | NSW | 2020-11-15 |
| EPI_ISL_2839202 | HRSV/A/Australia/WM3281795/2020 | NSW | 2020-11-23 |
| EPI_ISL_2839205 | HRSV/A/Australia/WM3301586/2020 | NSW | 2020-11-25 |
| EPI_ISL_2839204 | HRSV/A/Australia/WM3151215/2020 | NSW | 2020-11-10 |
| EPI_ISL_2839206 | HRSV/A/Australia/WM3150792/2020 | NSW | 2020-11-10 |
| EPI_ISL_2839419 | HRSV/A/Australia/WM3151529/2020 | NSW | 2020-11-10 |
| EPI_ISL_2839200 | HRSV/A/Australia/WM3210899/2020 | NSW | 2020-11-16 |
| EPI_ISL_2839188 | HRSV/A/Australia/WM3073568/2020 | NSW | 2020-11-02 |
| EPI_ISL_2839242 | HRSV/A/Australia/WM3301231/2020 | NSW | 2020-11-25 |
| EPI_ISL_2839198 | HRSV/A/Australia/WM3303286/2020 | NSW | 2020-11-25 |
| EPI_ISL_2839207 | HRSV/A/Australia/WM3145009/2020 | NSW | 2020-11-09 |
| EPI_ISL_2839211 | HRSV/A/Australia/WM3291444/2020 | NSW | 2020-11-24 |
| EPI_ISL_2839250 | HRSV/A/Australia/WM3220266/2020 | NSW | 2020-11-17 |
| EPI_ISL_2839248 | HRSV/A/Australia/WM3171598/2020 | NSW | 2020-11-12 |
| EPI_ISL_2839221 | HRSV/A/Australia/WM3142234/2020 | NSW | 2020-11-09 |
| EPI_ISL_2839362 | HRSV/A/Australia/WM3152673/2020 | NSW | 2020-11-10 |
| EPI_ISL_2839360 | HRSV/A/Australia/WM3341608/2020 | NSW | 2020-11-29 |
| EPI_ISL_2839326 | HRSV/A/Australia/WM3114268/2020 | NSW | 2020-11-06 |
| EPI_ISL_2839322 | HRSV/A/Australia/WM3293488/2020 | NSW | 2020-11-24 |
| EPI_ISL_2839324 | HRSV/A/Australia/WM3341974/2020 | NSW | 2020-11-29 |
| EPI_ISL_2839189 | HRSV/A/Australia/WM3080072/2020 | NSW | 2020-11-03 |
| EPI_ISL_2839332 | HRSV/A/Australia/WM3111229/2020 | NSW | 2020-11-06 |
| EPI_ISL_2839285 | HRSV/A/Australia/WM3300592/2020 | NSW | 2020-11-25 |
| EPI_ISL_2839339 | HRSV/A/Australia/WM3281572/2020 | NSW | 2020-11-23 |
| EPI_ISL_2839328 | HRSV/A/Australia/WM3160376/2020 | NSW | 2020-11-11 |
| EPI_ISL_2839334 | HRSV/A/Australia/WM3152749/2020 | NSW | 2020-11-10 |
| EPI_ISL_2839226 | HRSV/A/Australia/WM3253207/2020 | NSW | 2020-11-20 |
| EPI_ISL_2839187 | HRSV/A/Australia/WM3071478/2020 | NSW | 2020-11-02 |
| EPI_ISL_2839186 | HRSV/A/Australia/WM3071531/2020 | NSW | 2020-11-02 |
| EPI_ISL_2839246 | HRSV/A/Australia/WM3180542/2020 | NSW | 2020-11-13 |
| EPI_ISL_2839343 | HRSV/A/Australia/WM3221870/2020 | NSW | 2020-11-17 |

| | | | |
|---|---|---|---|
| EPI_ISL_2839320 | HRSV/A/Australia/WM3084781/2020 | NSW | 2020-11-03 |
| EPI_ISL_2839236 | HRSV/A/Australia/WM3212551/2020 | NSW | 2020-11-16 |
| EPI_ISL_2839318 | HRSV/A/Australia/WM3100311/2020 | NSW | 2020-11-05 |
| EPI_ISL_2839244 | HRSV/A/Australia/WM3302850/2020 | NSW | 2020-11-25 |
| EPI_ISL_2839283 | HRSV/A/Australia/WM3322984/2020 | NSW | 2020-11-27 |
| EPI_ISL_2839341 | HRSV/A/Australia/WM3101273/2020 | NSW | 2020-11-05 |
| EPI_ISL_2839345 | HRSV/A/Australia/WM3292889/2020 | NSW | 2020-11-24 |
| EPI_ISL_2839316 | HRSV/A/Australia/WM3301616/2020 | NSW | 2020-11-25 |
| EPI_ISL_2839196 | HRSV/A/Australia/WM3151688/2020 | NSW | 2020-11-10 |
| EPI_ISL_2839254 | HRSV/A/Australia/WM3081844/2020 | NSW | 2020-11-03 |
| EPI_ISL_2839260 | HRSV/A/Australia/WM3060152/2020 | NSW | 2020-11-01 |
| EPI_ISL_2839349 | HRSV/A/Australia/WM3324484/2020 | NSW | 2020-11-27 |
| EPI_ISL_2839220 | HRSV/A/Australia/WM3201552/2020 | NSW | 2020-11-15 |
| EPI_ISL_2839269 | HRSV/A/Australia/WM3280473/2020 | NSW | 2020-11-23 |
| EPI_ISL_2839271 | HRSV/A/Australia/WM3341337/2020 | NSW | 2020-11-29 |
| EPI_ISL_2839210 | HRSV/A/Australia/WM3321046/2020 | NSW | 2020-11-27 |
| EPI_ISL_2839222 | HRSV/A/Australia/WM3180278/2020 | NSW | 2020-11-13 |
| EPI_ISL_2839225 | HRSV/A/Australia/WM3301224/2020 | NSW | 2020-11-25 |
| EPI_ISL_2839209 | HRSV/A/Australia/WM3231345/2020 | NSW | 2020-11-18 |
| EPI_ISL_2839203 | HRSV/A/Australia/WM3321824/2020 | NSW | 2020-11-27 |
| EPI_ISL_2839201 | HRSV/A/Australia/WM3131917/2020 | NSW | 2020-11-08 |
| EPI_ISL_2839330 | HRSV/A/Australia/WM3331626/2020 | NSW | 2020-11-28 |
| EPI_ISL_2839420 | HRSV/A/Australia/WM3174399/2020 | NSW | 2020-11-12 |
| EPI_ISL_2839313 | HRSV/A/Australia/WM3291066/2020 | NSW | 2020-11-24 |
| EPI_ISL_2839239 | HRSV/A/Australia/WM3141851/2020 | NSW | 2020-11-09 |
| EPI_ISL_2839224 | HRSV/A/Australia/WM3202528/2020 | NSW | 2020-11-15 |
| EPI_ISL_2839219 | HRSV/A/Australia/WM3151206/2020 | NSW | 2020-11-10 |
| EPI_ISL_2839223 | HRSV/A/Australia/WM3323284/2020 | NSW | 2020-11-27 |
| EPI_ISL_2839262 | HRSV/A/Australia/WM3312926/2020 | NSW | 2020-11-26 |
| EPI_ISL_2839428 | HRSV/A/Australia/WM3291678/2020 | NSW | 2020-11-24 |
| EPI_ISL_2839358 | HRSV/A/Australia/WM3101170/2020 | NSW | 2020-11-05 |
| EPI_ISL_2839337 | HRSV/A/Australia/WM3151664/2020 | NSW | 2020-11-10 |
| EPI_ISL_2839191 | HRSV/A/Australia/WM3151642/2020 | NSW | 2020-11-10 |
| EPI_ISL_2839273 | HRSV/A/Australia/WM3351169/2020 | NSW | 2020-11-30 |
| EPI_ISL_2839292 | HRSV/A/Australia/WM3280848/2020 | NSW | 2020-11-23 |
| EPI_ISL_2839347 | HRSV/A/Australia/WM3312123/2020 | NSW | 2020-11-26 |
| EPI_ISL_2839258 | HRSV/A/Australia/WM3221617/2020 | NSW | 2020-11-17 |
| EPI_ISL_2839281 | HRSV/A/Australia/WM3171321/2020 | NSW | 2020-11-12 |
| EPI_ISL_2839457 | HRSV/B/Australia/WM0634962/2020 | NSW | 2020-03-03 |
| EPI_ISL_2839452 | HRSV/B/Australia/WM0742913/2020 | NSW | 2020-03-14 |
| EPI_ISL_2839416 | HRSV/B/Australia/WM3181191/2020 | NSW | 2020-11-13 |
| EPI_ISL_2839374 | HRSV/B/Australia/WM3181190/2020 | NSW | 2020-11-13 |

| | | | |
|---|---|---|---|
| EPI_ISL_2839435 | HRSV/B/Australia/WM0711659/2020 | NSW | 2020-03-11 |
| EPI_ISL_2839185 | HRSV/B/Australia/WM0663762/2020 | NSW | 2020-03-06 |
| EPI_ISL_2839393 | HRSV/B/Australia/WM0715241/2020 | NSW | 2020-03-11 |
| EPI_ISL_2839437 | HRSV/B/Australia/WM0331623/2020 | NSW | 2020-02-02 |
| EPI_ISL_2839394 | HRSV/B/Australia/WM0573055/2020 | NSW | 2020-02-26 |
| EPI_ISL_2839454 | HRSV/B/Australia/WM0636501/2020 | NSW | 2020-03-03 |
| EPI_ISL_2839180 | HRSV/B/Australia/WM0531461/2020 | NSW | 2020-02-22 |
| EPI_ISL_2839368 | HRSV/B/Australia/WM1603138/2020 | NSW | 2020-06-08 |
| EPI_ISL_2839417 | HRSV/B/Australia/WM0322845/2020 | NSW | 2020-02-01 |
| EPI_ISL_2839436 | HRSV/B/Australia/WM0543490/2020 | NSW | 2020-02-23 |
| EPI_ISL_2839193 | HRSV/B/Australia/WM0420543/2020 | NSW | 2020-02-11 |
| EPI_ISL_2839366 | HRSV/B/Australia/WM0421262/2020 | NSW | 2020-02-11 |
| EPI_ISL_2839442 | HRSV/B/Australia/WM0653812/2020 | NSW | 2020-03-05 |
| EPI_ISL_2839449 | HRSV/B/Australia/WM0782450/2020 | NSW | 2020-03-18 |
| EPI_ISL_2543785 | hRSV/A/Australia/VIC-RCH009/2021 | VIC | 2021-02-07 |
| EPI_ISL_2543770 | hRSV/A/Australia/VIC-MMC035/2020 | VIC | 2020-12-22 |
| EPI_ISL_2543786 | hRSV/A/Australia/VIC-RCH010/2021 | VIC | 2021-02-06 |
| EPI_ISL_2543771 | hRSV/A/Australia/VIC-MMC038/2020 | VIC | 2020-12-23 |
| EPI_ISL_2543772 | hRSV/A/Australia/VIC-MMC039/2020 | VIC | 2020-12-23 |
| EPI_ISL_2543784 | hRSV/A/Australia/VIC-RCH008/2021 | VIC | 2021-01-31 |
| EPI_ISL_2543764 | hRSV/A/Australia/VIC-MMC007/2021 | VIC | 2021-01-02 |
| EPI_ISL_1653939 | hRSV/A/Australia/VIC-RCH001/2021 | VIC | 2021-01-02 |
| EPI_ISL_2543781 | hRSV/A/Australia/VIC-MMC099/2021 | VIC | 2021-01-18 |
| EPI_ISL_1653942 | hRSV/A/Australia/VIC-RCH002/2021 | VIC | 2021-01-03 |
| EPI_ISL_2543769 | hRSV/A/Australia/VIC-MMC033/2020 | VIC | 2020-12-21 |
| EPI_ISL_2543800 | hRSV/A/Australia/VIC-RCH035/2021 | VIC | 2021-02-28 |
| EPI_ISL_2543765 | hRSV/A/Australia/VIC-MMC014/2020 | VIC | 2020-12-19 |
| EPI_ISL_2543774 | hRSV/A/Australia/VIC-MMC053/2020 | VIC | 2020-12-30 |
| EPI_ISL_1653941 | hRSV/A/Australia/VIC-RCH002/2020 | VIC | 2020-12-20 |
| EPI_ISL_2543787 | hRSV/A/Australia/VIC-RCH011/2021 | VIC | 2021-02-06 |
| EPI_ISL_2543807 | hRSV/A/Australia/VIC-RCH051/2021 | VIC | 2021-03-08 |
| EPI_ISL_1653944 | hRSV/A/Australia/VIC-VIDRL003/2020 | VIC | 2020-12-28 |
| EPI_ISL_2543797 | hRSV/A/Australia/VIC-RCH030/2021 | VIC | 2021-02-22 |
| EPI_ISL_2543789 | hRSV/A/Australia/VIC-RCH013/2021 | VIC | 2021-02-12 |
| EPI_ISL_2543780 | hRSV/A/Australia/VIC-MMC065/2021 | VIC | 2021-01-12 |
| EPI_ISL_2543805 | hRSV/A/Australia/VIC-RCH048/2021 | VIC | 2021-03-07 |
| EPI_ISL_1653948 | hRSV/A/Australia/VIC-VIDRL005/2020 | VIC | 2020-12-29 |
| EPI_ISL_2811729 | hRSV/A/Australia/VIC-RCH007/2020 | VIC | 2020-11-26 |
| EPI_ISL_1653946 | hRSV/A/Australia/VIC-VIDRL004/2020 | VIC | 2020-12-29 |
| EPI_ISL_2543795 | hRSV/A/Australia/VIC-RCH022/2021 | VIC | 2021-02-15 |
| EPI_ISL_2543778 | hRSV/A/Australia/VIC-MMC064/2020 | VIC | 2020-12-31 |
| EPI_ISL_2543766 | hRSV/A/Australia/VIC-MMC024/2020 | VIC | 2020-12-21 |

| | | | |
|---|---|---|---|
| EPI_ISL_2543804 | hRSV/A/Australia/VIC-RCH047/2021 | VIC | 2021-03-04 |
| EPI_ISL_2543790 | hRSV/A/Australia/VIC-RCH016/2021 | VIC | 2021-02-11 |
| EPI_ISL_2543767 | hRSV/A/Australia/VIC-MMC027/2020 | VIC | 2020-12-21 |
| EPI_ISL_2543796 | hRSV/A/Australia/VIC-RCH026/2021 | VIC | 2021-02-20 |
| EPI_ISL_2543783 | hRSV/A/Australia/VIC-RCH007/2021 | VIC | 2021-01-16 |
| EPI_ISL_2543798 | hRSV/A/Australia/VIC-RCH031/2021 | VIC | 2021-02-27 |
| EPI_ISL_2543775 | hRSV/A/Australia/VIC-MMC054/2020 | VIC | 2020-12-30 |
| EPI_ISL_2543762 | hRSV/A/Australia/VIC-MMC073/2021 | VIC | 2021-01-13 |
| EPI_ISL_1653938 | hRSV/A/Australia/VIC-VIDRL001/2020 | VIC | 2020-12-22 |
| EPI_ISL_2543777 | hRSV/A/Australia/MMC055/2021 | VIC | 2021-01-10 |
| EPI_ISL_2543773 | hRSV/A/Australia/MMC045/2020 | VIC | 2020-12-27 |
| EPI_ISL_2543850 | hRSV/B/Australia/VIC-RCH053/2021 | VIC | 2021-03-07 |
| EPI_ISL_2543763 | hRSV/B/Australia/VIC-RCH059/2021 | VIC | 2021-02-22 |
| EPI_ISL_2543852 | hRSV/B/Australia/VIC-RCH058/2021 | VIC | 2021-02-21 |
| EPI_ISL_2543853 | hRSV/B/Australia/VIC-RCH060/2021 | VIC | 2021-02-22 |
| EPI_ISL_2543849 | hRSV/B/Australia/VIC-RCH025/2021 | VIC | 2021-02-20 |
| EPI_ISL_2543851 | hRSV/B/Australia/VIC-RCH057/2021 | VIC | 2021-03-06 |
| EPI_ISL_2839418 | hRSV/A/Australia/1003117/2020 | WA | 2020-11-10 |
| EPI_ISL_2839431 | hRSV/A/Australia/2002690/2020 | WA | 2020-11-22 |
| EPI_ISL_2839432 | hRSV/A/Australia/5584/2020 | WA | 2020-11-30 |
| EPI_ISL_2839411 | hRSV/A/Australia/1008347/2021 | WA | 2021-02-01 |
| EPI_ISL_2839447 | hRSV/A/Australia/6002331/2020 | WA | 2020-12-06 |
| EPI_ISL_2839439 | hRSV/A/Australia/3012220/2020 | WA | 2020-12-03 |
| EPI_ISL_2839444 | hRSV/A/Australia/1002541/2021 | WA | 2021-01-01 |
| EPI_ISL_2839429 | hRSV/A/Australia/5003133/2021 | WA | 2021-01-04 |
| EPI_ISL_2839174 | hRSV/A/Australia/3160/2020 | WA | 2020-11-30 |
| EPI_ISL_2839433 | hRSV/A/Australia/8003496/2021 | WA | 2021-01-17 |
| EPI_ISL_2839171 | hRSV/A/Australia/3010918/2021 | WA | 2021-02-03 |
| EPI_ISL_2839443 | hRSV/A/Australia/3904/2020 | WA | 2020-12-10 |
| EPI_ISL_2839194 | hRSV/A/Australia/4010993/2020 | WA | 2020-12-04 |
| EPI_ISL_2839199 | hRSV/A/Australia/1643/2021 | WA | 2021-01-10 |
| EPI_ISL_2839445 | hRSV/A/Australia/3002027/2021 | WA | 2021-01-22 |
| EPI_ISL_2839438 | hRSV/A/Australia/1011044/2020 | WA | 2020-12-01 |
| EPI_ISL_2839423 | hRSV/A/Australia/3004170/2021 | WA | 2021-01-21 |
| EPI_ISL_2839446 | hRSV/A/Australia/4004352/2020 | WA | 2020-11-14 |
| EPI_ISL_2839441 | hRSV/A/Australia/6003904/2020 | WA | 2020-12-06 |
| EPI_ISL_2839398 | hRSV/A/Australia/3073/2020 | WA | 2020-11-20 |
| EPI_ISL_2839383 | hRSV/A/Australia/3002009/2020 | WA | 2020-12-12 |
| EPI_ISL_2839378 | hRSV/A/Australia/5005469/2021 | WA | 2021-01-05 |
| EPI_ISL_2839405 | hRSV/A/Australia/4009301/2020 | WA | 2020-12-04 |
| EPI_ISL_2839184 | hRSV/A/Australia/1007437/2021 | WA | 2021-02-01 |
| EPI_ISL_2839410 | hRSV/A/Australia/8010547/2020 | WA | 2020-11-18 |

| | | | |
|---|---|---|---|
| EPI_ISL_2839388 | hRSV/A/Australia/3002022/2020 | WA | 2020-12-12 |
| EPI_ISL_2839390 | hRSV/A/Australia/6009368/2020 | WA | 2020-11-26 |
| EPI_ISL_2839392 | hRSV/A/Australia/9002215/2020 | WA | 2020-11-19 |
| EPI_ISL_2839407 | hRSV/A/Australia/9002436/2020 | WA | 2020-11-28 |
| EPI_ISL_2839406 | hRSV/A/Australia/6003147/2020 | WA | 2020-12-06 |
| EPI_ISL_2839409 | hRSV/A/Australia/7003912/2020 | WA | 2020-11-17 |
| EPI_ISL_2839391 | hRSV/A/Australia/7007045/2020 | WA | 2020-11-27 |
| EPI_ISL_2839415 | hRSV/A/Australia/8002008/2020 | WA | 2020-11-27 |
| EPI_ISL_2839395 | hRSV/A/Australia/1013898/2021 | WA | 2021-02-01 |
| EPI_ISL_2839192 | hRSV/A/Australia/7006894/2020 | WA | 2020-12-07 |
| EPI_ISL_2839421 | hRSV/A/Australia/8006798/2021 | WA | 2021-01-28 |
| EPI_ISL_2839440 | hRSV/A/Australia/4011388/2020 | WA | 2020-11-04 |
| EPI_ISL_2839448 | hRSV/A/Australia/9005740/2020 | WA | 2020-10-29 |
| EPI_ISL_2839430 | hRSV/A/Australia/3001519/2020 | WA | 2020-11-03 |
| EPI_ISL_2839414 | hRSV/A/Australia/8001840/2020 | WA | 2020-10-08 |
| EPI_ISL_2839387 | hRSV/A/Australia/1003657/2020 | WA | 2020-11-01 |
| EPI_ISL_2839412 | hRSV/A/Australia/7006882/2020 | WA | 2020-11-27 |
| EPI_ISL_2839389 | hRSV/A/Australia/4005237/2020 | WA | 2020-10-13 |
| EPI_ISL_2835616 | hRSV/A/Australia/1514/2021 | WA | 2021-01-18 |
| EPI_ISL_2839179 | hRSV/A/Australia/1006644/2020 | WA | 2020-11-21 |
| EPI_ISL_2839195 | hRSV/A/Australia/8005369/2021 | WA | 2021-01-18 |
| EPI_ISL_2839176 | hRSV/A/Australia/6002591/2021 | WA | 2021-01-25 |
| EPI_ISL_2839381 | hRSV/A/Australia/2002105/2020 | WA | 2020-11-22 |
| EPI_ISL_2839177 | hRSV/A/Australia/6002451/2021 | WA | 2021-01-05 |
| EPI_ISL_2839364 | hRSV/A/Australia/9005762/2020 | WA | 2020-10-19 |
| EPI_ISL_2839172 | hRSV/A/Australia/9005783/2020 | WA | 2020-10-19 |
| EPI_ISL_2839399 | hRSV/A/Australia/3471/2020 | WA | 2020-10-20 |
| EPI_ISL_2839413 | hRSV/A/Australia/7494P/2020 | WA | 2020-09-03 |
| EPI_ISL_2839396 | hRSV/A/Australia/2010230/2020 | WA | 2020-11-12 |
| EPI_ISL_2839170 | hRSV/A/Australia/6545C/2020 | WA | 2020-08-20 |
| EPI_ISL_2839386 | hRSV/A/Australia/2051/2021 | WA | 2021-01-29 |
| EPI_ISL_2839370 | hRSV/A/Australia/1004203/2020 | WA | 2020-11-30 |
| EPI_ISL_2839183 | hRSV/A/Australia/6492G/2020 | WA | 2020-08-20 |
| EPI_ISL_2839385 | hRSV/A/Australia/7680J/2020 | WA | 2020-08-23 |
| EPI_ISL_2839182 | hRSV/A/Australia/7199H/2020 | WA | 2020-08-31 |
| EPI_ISL_2839175 | hRSV/A/Australia/6992X/2020 | WA | 2020-08-28 |
| EPI_ISL_2839402 | hRSV/A/Australia/3004/2021 | WA | 2021-01-09 |
| EPI_ISL_2839404 | hRSV/A/Australia/3002687/2021 | WA | 2021-01-01 |
| EPI_ISL_2839403 | hRSV/A/Australia/1010209/2021 | WA | 2021-01-11 |
| EPI_ISL_2839400 | hRSV/A/Australia/3002650/2021 | WA | 2021-01-01 |
| EPI_ISL_2839401 | hRSV/A/Australia/5009934/2021 | WA | 2021-01-04 |
| EPI_ISL_2839372 | hRSV/A/Australia/8006635/2020 | WA | 2020-11-28 |

| | | | |
|---|---|---|---|
| EPI_ISL_2839397 | hRSV/A/Australia/9009560/2021 | WA | 2021-01-19 |
| EPI_ISL_2839173 | hRSV/A/Australia/6012043/2021 | WA | 2021-01-06 |
| EPI_ISL_2839379 | hRSV/A/Australia/2003588/2020 | WA | 2020-12-12 |
| EPI_ISL_2839384 | hRSV/A/Australia/8001314/2020 | WA | 2020-11-27 |
| EPI_ISL_2839408 | hRSV/A/Australia/2001479/2020 | WA | 2020-10-21 |
| EPI_ISL_2839181 | hRSV/A/Australia/5516M/2020 | WA | 2020-07-27 |
| EPI_ISL_2839376 | hRSV/B/Australia/7527H/2019 | WA | 2019 |
| EPI_ISL_2839453 | hRSV/B/Australia/9391H/2019 | WA | 2019 |

*Locations include the Australian Capital Territory (ACT), New South Wales (NSW), Western Australia (WA) and Victoria (VIC).
